# Plasma proteomics reveals continuous molecular heterogeneity rather than discrete subtypes in Alzheimer’s disease

**DOI:** 10.64898/2026.07.04.26357296

**Authors:** Junyoung Park, Yann Le Guen

## Abstract

**INTRODUCTION:** Alzheimer’s disease is clinically and biologically heterogeneous. We asked whether plasma proteomics separates patients into discrete molecular subtypes or instead reflects continuous biological variation.

**METHODS:** We studied 5,895 Global Neurodegeneration Proteomics Consortium (GNPC) participants with Alzheimer’s disease or mild cognitive impairment using protein coexpression networks, clustering, and continuous molecular-axis analysis. External analyses used Stanford Alzheimer’s Disease Research Center (ADRC) biomarker/imaging data and UK Biobank proteomics.

**RESULTS:** Four continuous axes captured 81.5% of module-level proteomic variation. Although a two-cluster solution was reproducible, separation was weak and added little clinical information beyond the continuous axes. Stanford ADRC analyses showed selected fluid biomarker associations, but imaging and PET results did not provide consistent support. In UK Biobank, projected axes were more strongly related to *APOE* genotype and systemic hematologic, renal, lipid, inflammatory, and hepatic traits than to clear dementia-risk replication.

**DISCUSSION:** Plasma proteomics did not support robust Alzheimer’s disease subtypes. Continuous molecular coordinates better describe plasma proteomic heterogeneity and may guide future biological stratification.

**Research in Context:** *Systematic Review:* Prior work has established that Alzheimer’s disease is biologically heterogeneous. Neuropathological and imaging studies have described hippocampal-sparing, limbic-predominant, typical, cortical, or subcortical patterns, while transcriptomic and CSF proteomic studies have reported molecular subtypes with distinct cellular, immune, vascular, synaptic, or metabolic signatures. Large-scale plasma proteomic studies and biobank resources show that circulating proteins capture dementia-related and systemic biology, but plasma-based subtype claims require caution because circulating proteins may reflect both central nervous system-linked and peripheral processes. We therefore considered a robust plasma proteomic subtype claim to require reproducible clustering, clear separation, incremental clinical information beyond continuous dimensions, and directionally coherent support from external biomarkers, imaging, or independent cohorts.

*Interpretation:* In this study, a brain-region- and cell-system-inspired plasma proteomic subtyping hypothesis was not supported as a set of discrete molecular classes. In GNPC, the most reproducible clustering solution showed weak separation and provided little additional clinical information after accounting for four continuous module-derived axes. External analyses further supported a cautious interpretation: Stanford biomarker analyses showed partial but partly discordant associations, whereas UK Biobank Olink projections most strongly contextualized the axes through *APOE* genotype and systemic hematologic, renal, lipid, inflammatory, and hepatic traits rather than through unambiguous dementia-risk replication. Together, these results suggest that plasma proteomic heterogeneity in Alzheimer’s disease is better represented by continuous molecular coordinates than by hard subtype labels.

*Future Directions:* The present findings argue for a shift from categorical plasma-proteomic subtyping toward continuous molecular stratification in Alzheimer’s disease. Replication in independent Alzheimer’s disease and mild cognitive impairment cohorts, ideally with longitudinal follow-up and harmonized biomarker and imaging measures, will be necessary to determine whether these axes are stable and clinically useful. Integrating plasma proteomics with phosphorylated tau, neurodegeneration markers, amyloid and tau imaging, and structural MRI may help distinguish Alzheimer’s disease–linked variation from systemic physiology. Finally, brain-region and cell-type annotations should be interpreted as biological context rather than evidence of tissue origin unless supported by paired tissue, imaging, or mechanistic data.

## Background

Alzheimer’s disease (AD) is biologically and clinically heterogeneous [1,2]. Patients differ in age at onset, cognitive and behavioral presentation, biomarker burden, comorbidity profile, and rate of progression, even within established diagnostic categories. This heterogeneity complicates prognosis, trial enrichment, and interpretation of molecular biomarkers, and raises a central question for precision medicine: whether AD heterogeneity reflects discrete biological subtypes or continuous variation across overlapping disease processes.

A major source of biological motivation for AD subtyping comes from regional and cellular vulnerability. AD pathology and neurodegeneration do not affect all brain regions or cell populations equally; cortical, hippocampal, subcortical, glial, vascular, and synaptic systems show distinct patterns of susceptibility across disease stages and individuals [3]. Single-cell and multi-brain-region transcriptomic studies have further shown that AD-related molecular changes vary across cell classes and anatomical regions, providing a rationale for asking whether peripheral molecular measurements might capture coordinated biological states related to regional or cellular vulnerability [4,5].

Prior molecular studies have used brain transcriptomic and cerebrospinal-fluid proteomic data to define AD subtypes, providing important evidence that AD is not a single homogeneous biological process [6,7]. However, clustering-based subtype definitions require careful interpretation. Reproducible partitions can arise when individuals are distributed along continuous gradients, particularly when disease severity, *APOE*-related biology, or systemic physiology dominates the molecular signal. A robust subtype claim therefore requires more than stable clustering: it should show clear separation, balanced and interpretable groups, clinical or biomarker information beyond continuous dimensions, and coherent support in external data [8,9].

Plasma proteomics offers a scalable opportunity to study AD heterogeneity in large cohorts. Large plasma proteomic resources have demonstrated broad genetic and disease associations, showing that circulating proteins capture both disease-related and systemic biology [10,11]. At the same time, AD-focused proteomic studies indicate that brain, cerebrospinal fluid, and plasma capture overlapping but non-identical molecular information, requiring careful interpretation when relating plasma-derived patterns to underlying disease processes [12,13].

In this study, the original hypothesis was brain-region- and cell-system-inspired rather than brain-origin-assuming. We asked whether proteins annotated by cortical, subcortical, cerebellar, neural, glial, vascular, or systemic expression priors organize into plasma proteomic states that distinguish biologically meaningful AD and mild cognitive impairment (MCI) subgroups. These priors were derived from tissue and cell-type expression resources and AD cellular atlases, and were used to interpret emergent protein structure, not to define the network or infer the tissue source of circulating proteins [14–17].

To test this hypothesis, we used weighted protein coexpression modules as an intermediate representation of plasma proteomic structure [18]. Coexpression modules summarize coordinated protein variation without requiring prespecified diagnostic labels or regional categories. We then compared discrete clustering of module eigengenes with continuous decomposition of the same module space, allowing us to ask whether the data supported hard subtype labels or whether participant heterogeneity was better represented by continuous molecular coordinates linked to modules, proteins, clinical traits, and biological annotations.

External evaluation was matched to the level of claim. In GNPC, we tested whether clusters were stable, separated, clinically informative, and incremental beyond continuous axes. In Stanford ADRC, we evaluated whether frozen axes showed coherent associations with fluid biomarkers and neuroimaging traits in a clinically characterized cohort. In UK Biobank, we projected GNPC axes to Olink proteomics to assess cross-platform biological context, including dementia risk, *APOE*, cognition, MRI, and systemic phenotypes. Because UK Biobank used a different proteomic platform and a population-based design, it was treated as contextualization rather than direct replication of GNPC SomaScan subtypes.

We therefore tested whether brain-region- and cell-system-inspired plasma proteomic structure supports discrete AD/MCI subtypes or continuous molecular heterogeneity. The analysis was designed to retain a subtype interpretation only if discrete clusters showed clear separation, reproducibility, incremental clinical value, and directionally coherent external support. Otherwise, the more defensible representation would be continuous molecular coordinates that capture graded neural, vascular-glial, APOE/lipid, and systemic components of plasma proteomic heterogeneity.

## Methods

### Study design and cohorts

We conducted a multi-cohort plasma proteomic study to test whether a brain-region- and cell-system-inspired hypothesis of Alzheimer’s disease (AD) heterogeneity was better represented by discrete molecular subtypes or by continuous molecular axes. Regional and cellular expression priors motivated the biological question but did not constrain network construction, axis derivation, or cluster assignment.

The Global Neurodegeneration Proteomics Consortium (GNPC) SomaScan dataset served as the discovery cohort. Stanford Alzheimer’s Disease Research Center (ADRC) data were used for external association analyses with fluid biomarkers and neuroimaging traits. UK Biobank (UKB) Olink proteomics were used for population-based cross-platform contextualization. The overall analytic framework is shown in **Figure 1**. GNPC module discovery, axis definition, protein weights, and axis orientation were completed before external interpretation. Stanford and UKB analyses did not repeat WGCNA or subtype discovery, and UKB was not used to reproduce GNPC AD/MCI clusters.

**Figure 1.**
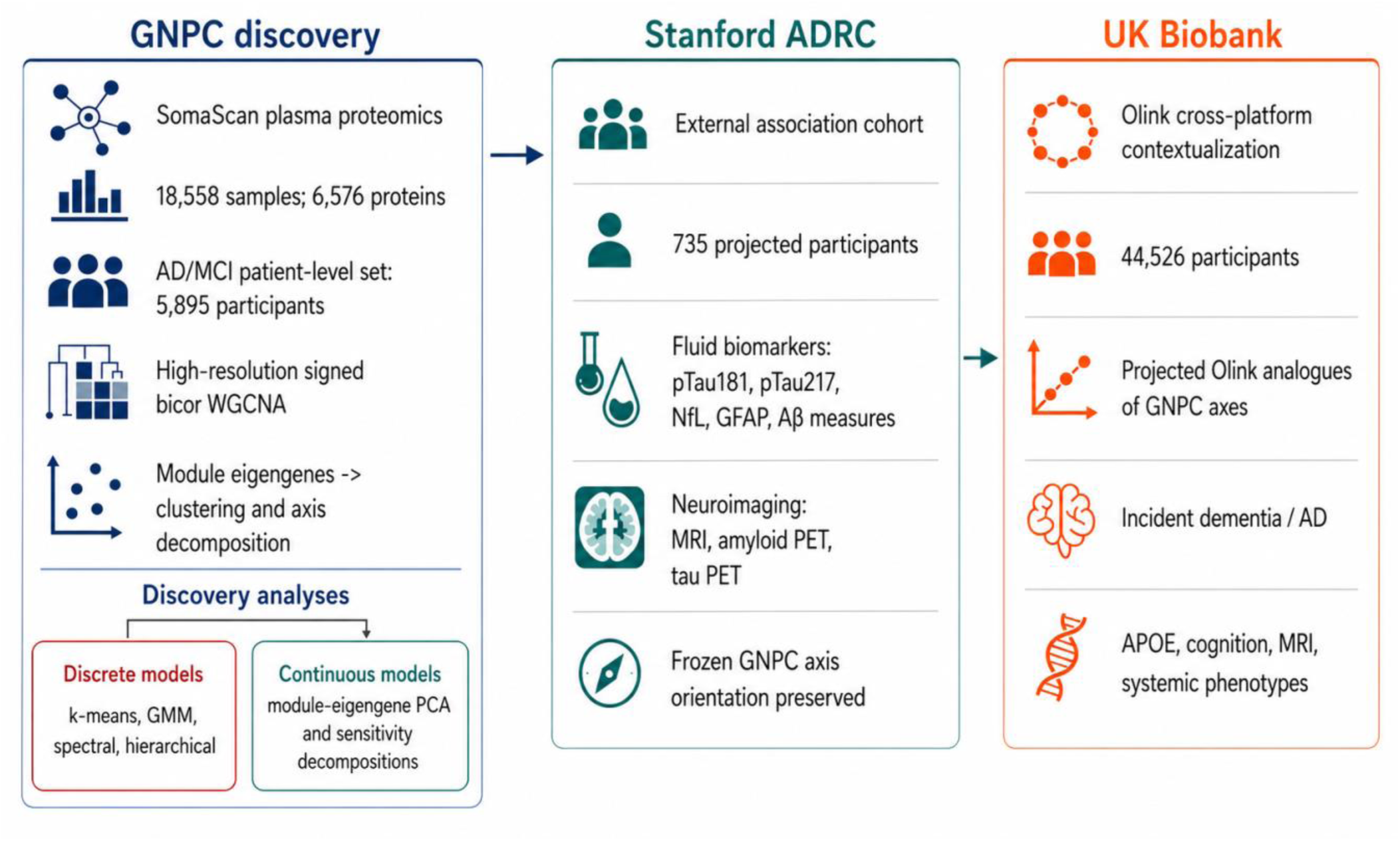
Analytic framework for testing plasma proteomic subtype structure. GNPC SomaScan data were used for discovery, including high-resolution signed biweight midcorrelation (bicor) WGCNA, patient-level module-eigengene analysis, discrete clustering models, and continuous axis decompositions. Frozen GNPC-derived axes were then evaluated in Stanford ADRC for external fluid biomarker and neuroimaging associations and in UK Biobank for Olink-based cross-platform contextualization. UK Biobank scores represent Olink analogues of GNPC protein contrasts, not direct SomaScan replication.

The GNPC all-sample network included 18,558 samples and 6,576 collapsed human proteins. The AD/MCI network included 6,598 samples, from which one record per participant was selected for patient-level decomposition, yielding 5,895 unique AD/MCI participants. The AD/MCI network included 3,182 AD-labeled and 3,416 MCI/SCD-labeled samples, with mean age 73.3 years; 4,733 samples had *APOE* dosage and 6,557 had computed Clinical Dementia Rating (CDR). Records with sex codes other than 1 or 2 were excluded. Cohort and sample characteristics are summarized in **Table S1**

The Stanford ADRC resource included 1,165 plasma samples from 735 participants. Within 365 days of plasma sampling, 823 samples matched fluid biomarker visits, 187 matched cortical-thickness MRI, 136 matched amyloid PET, and 27 matched tau PET. These counts describe available linked resources; model-specific sample sizes were determined by complete outcome and covariate data. The UKB Olink resource was generated by the UK Biobank Pharma Proteomics Project (UKB-PPP) using the Olink Explore 3072 platform on plasma samples from 54,219 UKB participants [19]. For the present analysis, we used the post-UKB-PPP quality-controlled and imputed baseline 3k Olink matrix generated under UKB application 45420, following the workflow we described in [20]. Briefly, the baseline post-QC resource consisted of 2,923 protein measurements from 53,018 samples. Samples with extensive missingness, proteins with >10% missingness, and samples with discordant reported and genetic sex were removed, resulting in 44,498 samples and 2,916 protein measurements before imputation. Missing values were imputed using a k-nearest-neighbor strategy after center-based train/test splitting and z-score normalization, with imputation performance assessed against a masked complete-case subset. The baseline matrix used for GNPC-axis projection in this study included 44,526 participants and 2,916 proteins after harmonization to the available analytic file.

### Network construction and axis derivation

Weighted gene coexpression network analysis (WGCNA) was used to summarize coordinated plasma protein variation [18]. Separate signed networks were constructed in all GNPC samples and in the AD/MCI subset using the biweight midcorrelation (“bicor”), a robust correlation measure less sensitive to outlying protein values. The primary networks used high-resolution module detection (deepSplit=4, minModuleSize=20, mergeCutHeight=0.10) to capture finer-grained coexpression structure. Age, sex, APOE, diagnosis, and CDR were not residualized before network discovery. Standard-resolution networks were retained as sensitivity scaffolds.

Module eigengenes summarized non-grey modules, and protein-module membership was quantified using kME. Principal component analysis of the nine non-grey AD/MCI module eigengenes defined the primary participant-level molecular axes. Axis signs were frozen after discovery to preserve positive and negative protein directions. Four axes were retained for interpretation and for testing whether discrete clusters added information beyond continuous molecular structure. Full-proteome principal components and alternative decompositions were evaluated as sensitivity representations. Primary module characteristics and frozen axis loadings are provided in **Table S2**.

GNPC axis-trait analyses used robust linear or logistic models, depending on the endpoint. Adjusted diagnosis models included age, valid sex, and *APOE* dosage. Adjusted CDR models additionally included diagnosis to distinguish within-diagnosis severity from case-control composition. Longitudinal models included baseline age, valid sex, *APOE* dosage, and baseline CDR when available. Marginal and adjusted estimates were retained side by side because covariate adjustment changed the biological question rather than simply refining precision. Sample sizes varied by endpoint according to outcome and covariate availability. False-discovery rate (FDR) was controlled using the Benjamini-Hochberg procedure [21].

### Discrete subtype tests

We evaluated whether the AD/MCI module eigengene space supported discrete subtypes. K-means, Gaussian-mixture, spectral, and hierarchical clustering were applied across multiple values of k. Clustering performance was assessed using separation, component size, information criteria where applicable, and repeated-subsample stability. Silhouette width was used to quantify separation, and adjusted Rand index was used to quantify assignment stability [8,9].

A reproducible partition was not considered sufficient evidence for a biological subtype. We therefore tested whether cluster assignment improved clinical, progression, conversion, or external biomarker models after accounting for age, sex, *APOE* dosage, and the four continuous molecular axes. Cluster solutions were interpreted as candidate subtypes only if they were stable, clearly separated, non-trivial in size, clinically informative beyond the axes, and directionally coherent in external analyses.

### Protein and RNA-derived prior annotation

Axes were interpreted using signed module loadings and individual protein loadings. Module assignment, kME, protein loading, and axis direction were reported separately to preserve the contrast structure of each axis.

RNA-derived priors were used for biological annotation, not for network construction or subtype assignment. Brain-region, tissue, body-cell-type, brain-cell-type, and AD cellular marker sets were derived from GTEx, the Human Protein Atlas, and SEA-AD resources [14–17,22]. Enrichment tests used measured assay genes as the background. Positive and negative ends of each axis were analyzed separately because each axis represents a signed molecular contrast. These analyses assessed compatibility with regional or cellular expression programs; they were not interpreted as evidence that circulating proteins originated from those tissues or cell types.

### Stanford ADRC biomarkers association analyses

Stanford SomaScan proteins were harmonized to the GNPC protein identifiers using the prespecified SomaScan crosswalk between versions, and participant scores were computed using the frozen GNPC protein weights and axis orientations. Plasma samples were linked to the nearest available fluid biomarker or imaging visit by participant and date or age. Outcomes included pTau181, pTau217, neurofilament light, GFAP, amyloid-beta measures, MRI cortical thickness, amyloid PET, and tau PET [23,24].

Standardized association models used available age, sex, diagnosis, *APOE* genotype dosages, and plasma-to-outcome interval as covariates. Exact formulas and sample sizes are reported in the supplementary tables. Fluid models included 398 participants for most biomarkers and 424 for pTau181. MRI-thickness models included 164 participants, amyloid PET models included 121, and tau PET models included 25.

Imaging variables were organized into global cortical, medial temporal, AD-signature, temporal, posterior/default-mode, frontal-parietal, subcortical, and cerebellar groupings where available. Because tau PET sample size was limited, tau PET estimates were considered exploratory. FDR was controlled within outcome modality.

### UK Biobank cross-platform projection and outcomes

GNPC axes were projected into UKB Olink proteomics to assess cross-platform biological context. Olink assays were mapped to GNPC proteins by gene symbol and standardized within matrix and visit. Duplicate Olink assays were averaged after standardization; highest-coverage assay selection was evaluated as a sensitivity analysis. Projected scores used normalized signed GNPC loadings. Axis 1 and Axis 4 met projectability criteria and were considered primary UKB projections. Axis 2 and Axis 3 had limited bidirectional coverage and were treated as exploratory. These projected scores are Olink analogues of the GNPC SomaScan axes, not direct replication of SomaScan measurements [10,11].

Incident dementia and AD analyses used attained age as the time scale, with delayed entry at the Olink sample [25]. Algorithmic fields defined all-cause dementia and AD event dates, and prevalent cases were excluded from the corresponding endpoint [26]. Participants were censored at death or a last-known-age aggregated from electronic health records and UK Biobank visits. The dementia risk set included 38,239 participants with 1,311 incident events, and the AD risk set included 38,269 participants with 693 incident events; median follow-up was 11.33 years. Base Cox models and APOE-adjusted Cox models were presented separately.

Cross-sectional UKB models adjusted for age, sex, ten genetic principal components, assessment center, Olink plate, and well. Prespecified outcome families included cognition, MRI, blood counts and platelets, inflammation, lipids, kidney function, and liver function. Neurobiomarker models were withheld when complete aligned data were insufficient. The UKB 1k Olink panel was used only for longitudinal sensitivity analyses. This earlier Olink platform measured approximately 1,500 proteins and was generated for 1,268 UKB participants who participated in the COVID-19 repeat imaging study. The available samples included the baseline visit, Instance 2 imaging visit, and Instance 3 repeat imaging visit. Because this dataset used a smaller earlier-version Olink panel and represented a repeat-imaging subset rather than an independent AD/MCI cohort, it was used to estimate across-visit stability of projected axis scores, not as independent replication or confirmation. Additional details on the UKB proteomics resource are provided by Sun et al. [19] and the UKB-PPP companion documentation.

Technical and biological adjustments were kept conceptually separate. Plate and well addressed available Olink processing structure, whereas *APOE*-adjusted incident models evaluated whether an axis proxy carried information conditional on genotype. Because *APOE* is a component and dominant correlate of Axis 4, *APOE* adjustment changes the estimand rather than simply removing confounding. Base and *APOE*-adjusted estimates were therefore presented together, and sign reversals were not interpreted as simple protective effects.

### Multiplicity and evidence tiers

Effect estimates were reported with standard errors or confidence intervals, nominal p values, analysis-specific sample sizes, and Benjamini-Hochberg q-values [21]. FDR was controlled within prespecified families, including clinical, progression, biomarker, imaging, systemic-trait, and RNA-prior analyses; global q values were also reported for UKB prior analyses.

Primary inference focused on the GNPC continuum-versus-cluster comparison, adjusted GNPC axis associations, Stanford modality-level external associations, and UKB Axis 1 and Axis 4 projections. Higher-k clusters, UKB Axis 2 and Axis 3 projections, individual imaging regions, tau PET, and source-specific prior findings were considered exploratory.

## Results

### Discrete clustering did not support robust AD/MCI plasma proteomic subtypes

We first tested whether the AD/MCI module eigengene space supported discrete participant groups. The selected k-means k=2 solution divided 5,895 participants into groups of 2,703 and 3,192. This partition was highly reproducible under repeated subsampling, with an adjusted Rand index of 0.975, but separation was weak, with a silhouette width of 0.251 (**Figure 2; Table S3**). In the two-dimensional Axis 1–Axis 2 projection, the clusters occupied opposing regions of a continuous distribution rather than distinct low-density-separated clouds.

**Figure 2.**
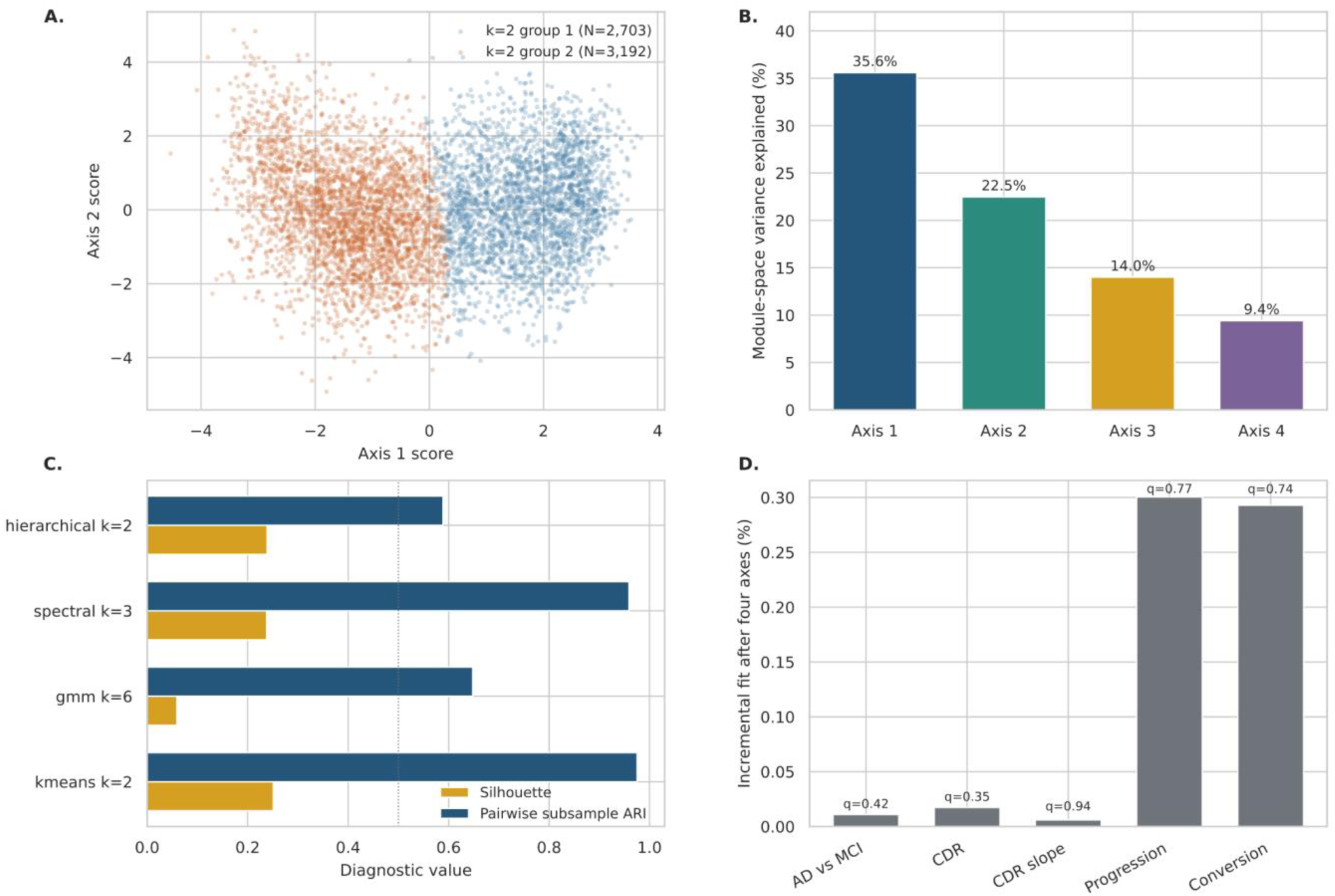
Discrete clustering and continuous-axis structure in GNPC AD/MCI plasma proteomics. (A) Patient-level projection of the first two module-derived axes, colored by the selected k-means k=2 assignment. The two groups were reproducible but occupied opposing regions of a dense continuous distribution rather than clearly separated clusters. (B) Percentage of AD/MCI module-space variance explained by the four retained continuous axes. (C) Cluster separation and stability for selected clustering configurations, summarized by silhouette width and pairwise subsample adjusted Rand index. (D) Incremental fit of the k=2 assignment after adjustment for the four continuous axes across GNPC clinical and progression outcomes. Together, these diagnostics indicated that clustering stability did not translate into strong separation or meaningful incremental clinical information.

Cluster assignment added little information after the four continuous axes. After adjustment for age, sex, *APOE* dosage, and the four axes, the k=2 assignment minimally changed model fit for AD versus MCI/SCD (change in pseudo-R²=0.00011; q=0.422) and CDR (change in R²=0.00017; q=0.351). It also did not add significant information for progression outcomes or Stanford biomarker associations. Thus, the most reproducible partition largely discretized a continuous module-space gradient rather than identifying a clinically distinct molecular state.

Higher-k solutions did not provide stronger subtype evidence. Spectral clustering with k=3 was stable (adjusted Rand index=0.959) but weakly separated (silhouette=0.238) and included a 434-person tail group. The Gaussian-mixture k=6 solution showed poor separation (silhouette=0.059), moderate stability (adjusted Rand index=0.648), and component sizes ranging from 506 to 1,660 participants. Although selected higher-k models captured small nonlinear differences in GNPC diagnosis or CDR, none improved Stanford biomarker models after accounting for the continuous axes. Because no clustering solution satisfied the prespecified criteria for robust subtypes, subsequent analyses used continuous axes as the primary participant-level representation. Expanded clustering diagnostics across k and clustering methods are shown in **Figure S1**.

### Four continuous axes captured most GNPC module-space variation

Principal component decomposition of the nine non-grey AD/MCI module eigengenes identified four continuous axes that explained 81.5% of module-space variance. Axes 1 through 4 explained 35.6%, 22.5%, 14.0%, and 9.4% of variance, respectively (**Figure 2**). Unlike cluster labels, these axes preserved participant ordering and retained bidirectional module and protein contributions. Sensitivity decompositions supported a low-dimensional continuous representation, while the module-derived axes preserved direct links to coexpression modules and individual proteins.

Axis 1 contrasted a turquoise systemic/scaffold-associated module with yellow and blue modules enriched for neural-guidance-related proteins (**Figure 3; Table S5**). The negative end included HSP90AA1, C11ORF68, MAPK14, NUDC, CRKL, and NAA10, whereas the positive end included GAS1, FLRT2, LRP10, EFNA5, EFNA2, and UNC5B. Higher Axis 1 scores were associated with AD versus MCI/SCD (adjusted beta=0.962; q=2.46×10^−123^), higher CDR (beta=0.095; q=1.54×10^−11^), and older age (beta=0.303; q=4.22×10^−61^), but not adjusted progression. We therefore interpreted Axis 1 as a severity/systemic scaffold–neural guidance contrast, without implying a brain-derived plasma signal.

**Figure 3.**
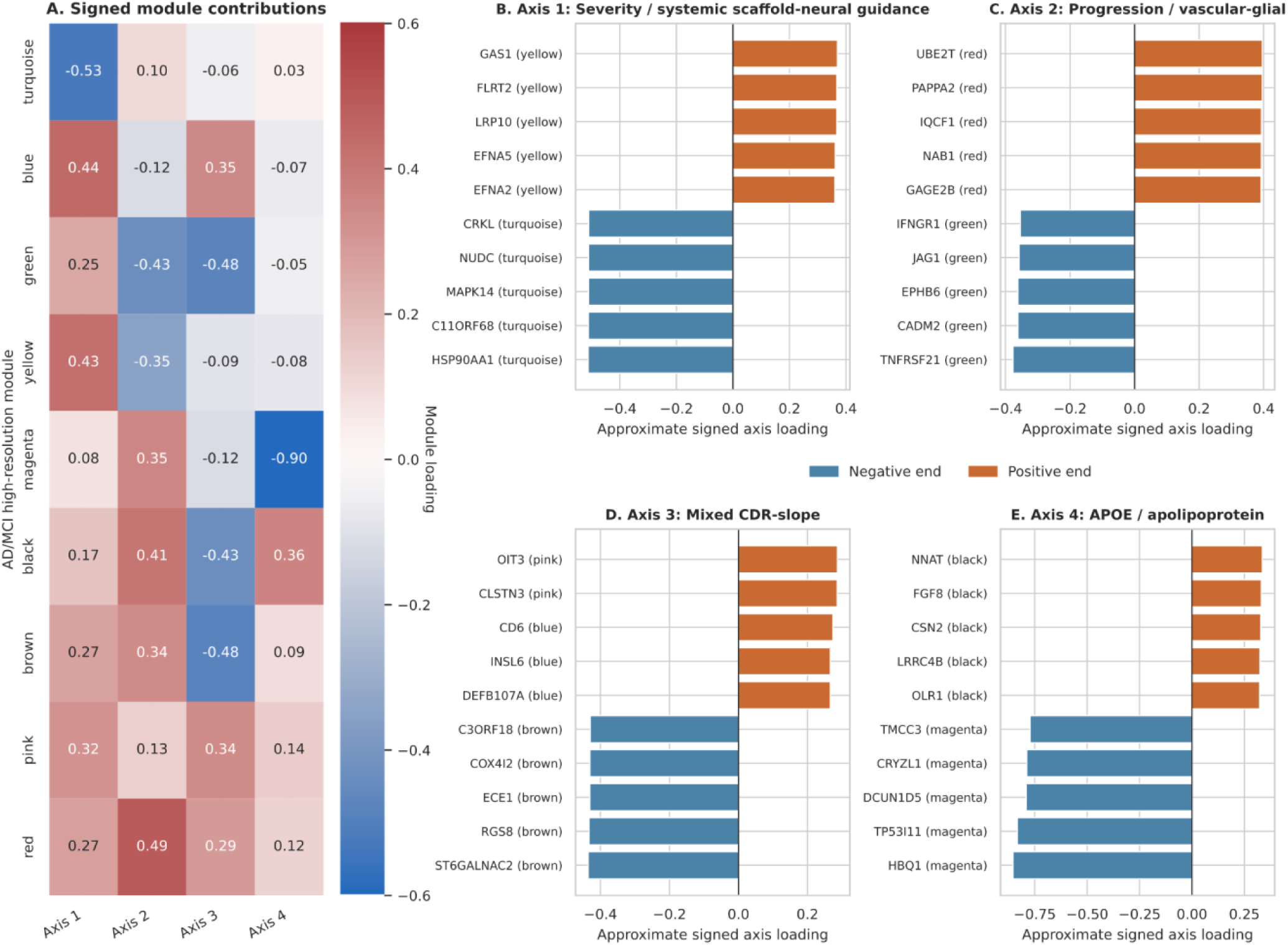
Module and protein interpretation of the four GNPC continuous axes. (A) Signed loadings of the nine non-grey AD/MCI high-resolution WGCNA modules on Axes 1–4. Positive and negative values indicate opposite ends of each molecular contrast. (B–E) Leading protein contributors at the positive and negative ends of each axis, with WGCNA module assignments shown in parentheses. Axis labels summarize the observed module, protein, and trait patterns: Axis 1, severity/systemic scaffold–neural guidance; Axis 2, progression/vascular-glial; Axis 3, mixed CDR-slope; and Axis 4, APOE/apolipoprotein. These labels are descriptive of plasma proteomic contrasts and do not imply tissue or cell origin.

Axis 2 contrasted red and black module states with green and yellow vascular- and glial-annotated modules. The negative end included TNFRSF21, CADM2, EPHB6, JAG1, IFNGR1, and ENG, whereas the positive end included UBE2T, PAPPA2, IQCF1, NAB1, GAGE2B, and PRLH. Axis 2 was associated with AD versus MCI/SCD (beta=0.394; q=8.29×10^−29^), CDR progression (beta=0.514; q=0.00645), and conversion to AD (beta=0.584; q=0.00674). These findings supported interpretation of Axis 2 as a progression/vascular-glial contrast, while recognizing the smaller longitudinal sample and limited cross-platform Olink coverage.

Axis 3 had a more mixed composition. Negative contributors included ST6GALNAC2, RGS8, ECE1, COX4I2, C3ORF18, and SEL1L2, whereas positive contributors included OIT3, CLSTN3, CD6, INSL6, DEFB107A, and ROBO3. Axis 3 was associated with AD versus MCI/SCD (beta=0.534; q=2.19×10^−57^) and lower subsequent CDR slope (beta=-0.139; q=0.0106), but its biological interpretation remained less specific than Axis 1 or Axis 2.

Axis 4 was dominated by the magenta APOE module and contrasted APOE, TP53I11, TMCC3, and related negative contributors with OLR1 and other positive contributors. Axis 4 showed a strong *APOE* ε4 dosage association (beta=0.222; q=5.84×10^−132^), supporting an APOE/apolipoprotein continuum rather than a discrete *APOE*-defined subtype. Expanded axis-level protein loading patterns are shown in **Figure S2**.

### RNA-derived priors provided biological annotation but not tissue-of-origin evidence

RNA-derived priors provided partial biological annotation of the GNPC axes (**Table S6**). Direction-specific foregrounds showed brain-focused annotations for Axis 1 and vascular/glial annotations for Axis 2, whereas Axis 3 lacked a direction-specific stringent signal. Systemic biology was also evident: the turquoise module was enriched for platelet/megakaryocyte markers, and the yellow module was enriched for monocyte, macrophage, and dendritic markers. No axis-level body-wide Human Protein Atlas enrichment survived the prespecified threshold.

At the protein level, annotations were heterogeneous within each axis. Proteins assigned to neural or brain-region expression programs occurred alongside proteins linked to platelet, immune, endothelial, metabolic, and broadly expressed cellular programs. This mixture was expected for plasma proteomics and argued against naming any entire axis after a single cell class or anatomical system. We therefore interpreted RNA-derived priors as biological context, not as evidence for the tissue or cellular origin of measured circulating proteins.

### Stanford external analyses provided partial and partly discordant support

After the GNPC structure was frozen, Stanford data were used to test whether axis directions were supported by fluid biomarkers and neuroimaging traits. Stanford analyses were not used to redefine modules, axes, or clusters.

Four Stanford fluid biomarker associations passed FDR correction (**Figure 4; Table S7**). Axis 1 was inversely associated with pTau181 (beta=-0.243; q=3.87×10^−6^), despite its positive association with AD diagnosis and CDR in GNPC. Axis 1 associations with pTau217 and neurofilament light were also negative but did not pass FDR correction, and Stanford diagnosis was null. The pTau181 association was therefore interpreted as directionally discordant external evidence rather than confirmation of a severity axis.

**Figure 4.**
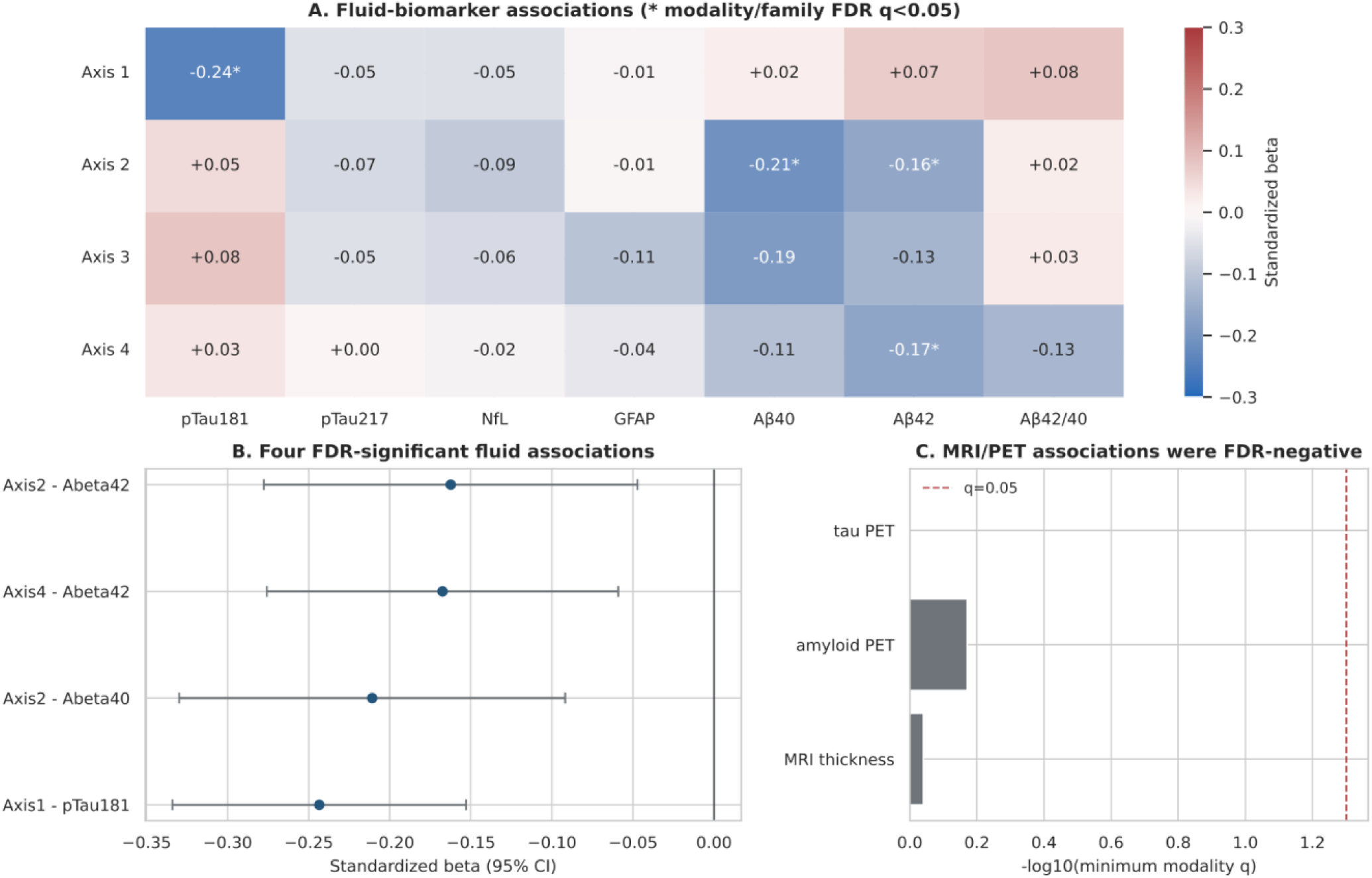
Stanford ADRC external biomarker and imaging associations of frozen GNPC axes. (A) Standardized associations between frozen GNPC-derived axes and Stanford fluid biomarkers. Asterisks denote associations passing modality/family false-discovery-rate correction at q<0.05. (B) Standardized effect estimates and 95% confidence intervals for the four FDR-significant fluid biomarker associations. (C) Minimum modality-level q values for MRI cortical thickness, amyloid PET, and tau PET associations; none passed FDR correction. Tau PET inference was limited by small sample size. Stanford analyses provided selected fluid-biomarker associations, including a directionally discordant Axis 1–pTau181 association, but did not provide coherent diagnostic, MRI, amyloid PET, or tau PET confirmation.

Axis 2 was inversely associated with amyloid-beta40 (beta=-0.211; q=0.00721) and amyloid-beta42 (beta=-0.162; q=0.0401). Its AD-versus-MCI association in Stanford was suggestive after FDR correction (q=0.065). Axis 4 was inversely associated with amyloid-beta42 (beta=-0.167; q=0.0229), but Stanford diagnosis was null. No axis association with MRI cortical thickness, amyloid PET, or tau PET passed modality-level FDR correction (**Figure S3**). The low tau PET sample size limited inference for that modality.

Overall, Stanford supplied selected fluid-biomarker associations but did not provide coherent diagnostic, imaging, or pathology-specific confirmation. The significant fluid results did not converge on a single external severity ordering, and the imaging null results prevented claims of region-specific or pathology-specific plasma axes. Sensitivity windows did not justify promoting nominal regional findings over the modality-level results.

### UK Biobank Olink projections contextualized axes primarily through systemic biology

UK Biobank was used to assess whether constrained Olink analogues of the GNPC axes retained biological and clinical context in a population cohort. It was not treated as a validation cohort for GNPC AD/MCI subtype assignments or as direct replication of SomaScan measurements. Interpretation therefore focused on projection coverage, direction, systemic correlates, and the limits of incident disease and prior-enrichment analyses.

Axis 1 and Axis 4 met primary Olink projectability criteria, with 20 and 23 mapped genes, respectively (**Figure 5; Figure S4; Table S8**). Axis 2 and Axis 3 had limited bidirectional coverage and were treated as exploratory proxies. Projected Axis 1 and Axis 4 score distributions in UKB are shown in **Figure S5**. Axis 4 aligned strongly with *APOE* genotype: each ε4 allele corresponded to a 0.174-SD higher score (q=1.4×10^−91^), whereas each ε2 allele corresponded to a 0.157-SD lower score (q=2.7×10^−42^).

**Figure 5.**
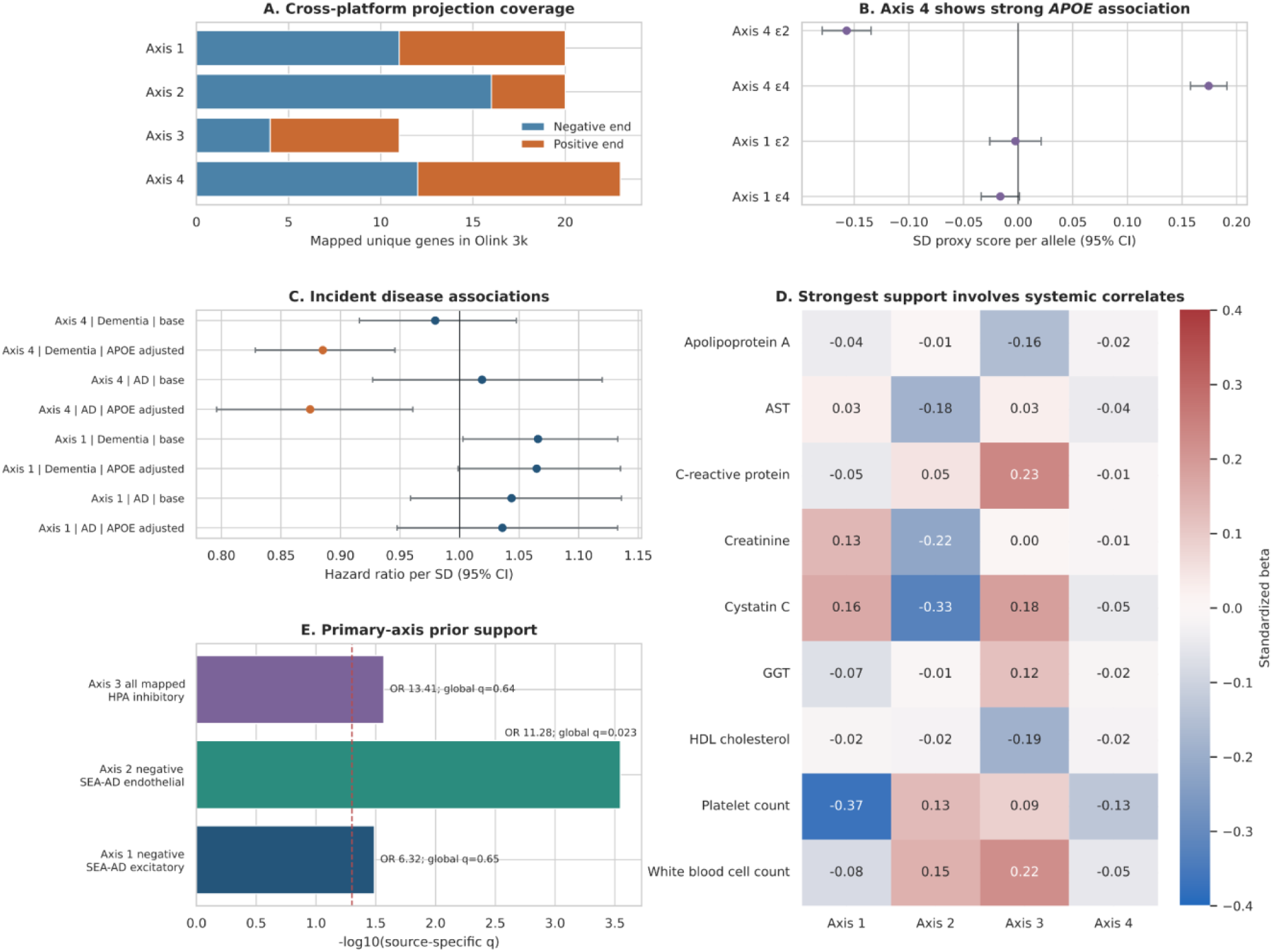
UK Biobank Olink cross-platform contextualization of GNPC-derived axes. (A) Number of mapped positive- and negative-end genes used to construct Olink analogues of the four GNPC axes. Axis 1 and Axis 4 were considered primary projections; Axis 2 and Axis 3 had limited bidirectional coverage and were treated as exploratory. (B) *APOE* ε2 and ε4 associations with primary Olink-projected axis scores. (C) Base and *APOE*-adjusted incident all-cause dementia and Alzheimer’s disease associations. These models did not provide unambiguous primary-axis dementia-risk evidence; Axis 4 estimates after *APOE* adjustment represent conditional/suppression contrasts rather than straightforward protection. (D) Representative systemic phenotype associations, highlighting the strongest UK Biobank context for the projected axes. (E) Selected RNA-derived prior enrichments for projected axes. Primary-axis prior support was limited, Axis 2 and Axis 3 findings were exploratory, and Axis 4 showed no significant set-level enrichment. UK Biobank analyses contextualized GNPC-derived protein contrasts across platforms but did not validate GNPC subtype assignments or establish tissue origin.

Incident disease associations were mixed and interpreted cautiously (**Table S9**). Axis 1 showed a nominal association with all-cause dementia in the base model (hazard ratio=1.066; 95% CI, 1.003–1.133; p=0.0405; within-model q=0.162) but was not associated with incident AD. Axis 4 was null in base dementia (hazard ratio=0.980; q=0.547) and AD (hazard ratio=1.019; q=0.931) models. After *APOE* genotype adjustment, Axis 4 estimates became inverse for dementia and AD. Because *APOE* is embedded in and strongly correlated with Axis 4, this reversal was interpreted as a conditional or suppression contrast rather than as evidence of protection or independent confirmation.

Systemic associations were stronger and more consistent than risk, cognition, or imaging evidence (**Figure 5; Figure S6; Table S10**). Axis 1 was associated with lower platelet count (beta=-0.370), higher creatinine (beta=0.134), higher cystatin C (beta=0.165), and lipid, C-reactive protein, blood-count, and liver traits. Axis 4 was associated with platelet, lymphocyte, lipid, kidney, and liver traits. Primary Axis 1 and Axis 4 MRI tests were FDR-negative. The exploratory Axis 3 proxy was associated with lower normalized total brain volume (beta=-0.042), greater white-matter hyperintensity burden (beta=0.039), and lower entorhinal thickness (beta=-0.036), but its cross-platform coverage was limited. Neurobiomarker models were withheld because complete aligned sample size was 126.

Cognitive associations for the primary Olink proxies were statistically detectable but small, approximately 0.01 to 0.03 SD per SD of proxy score. This magnitude contrasted with larger platelet and renal associations and reinforced the conclusion that the strongest UKB context was systemic. We did not build a multivariable dementia classifier or evaluate discrimination, calibration, or decision utility because the UKB objective was biological contextualization of the projected GNPC axes.

Cell-type prior enrichment of proteomics modules was limited (**Figure S7; Table S11**). The Axis 1 negative end retained GNPC-concordant SEA-AD excitatory-neuron enrichment (odds ratio=6.32; source-specific q=0.033), but the global q-value was 0.65 and GNPC positive-end cortex/cerebellum annotations were not retained. Axis 4 had no significant body or brain set-level enrichment. Exploratory Axis 2 showed endothelial enrichment (odds ratio=11.28; q=0.00028), and exploratory Axis 3 showed inhibitory-neuron enrichment (odds ratio=13.41; q=0.027). No primary axis had significant stringent Human Protein Atlas body-system enrichment. These results did not support a brain-specific plasma axis or tissue-origin interpretation.

Longitudinal sensitivity analyses in the nested 1k Olink panel included 1,219 participants with repeat scores. Intraclass correlations were moderate, ranging from 0.44 to 0.54, indicating both persistent between-person differences and within-person or measurement variation (**Figure S8**). These estimates supported limited temporal stability of the projected scores but did not constitute independent confirmation.

Taken together, UKB supported the existence of measurable cross-platform analogues and clarified that their strongest correlates were systemic. It did not provide unambiguous primary-axis dementia-risk evidence, primary-axis MRI support, or strong set-level neural enrichment. Axis 2 endothelial and Axis 3 MRI/inhibitory-neuron findings remain hypothesis-generating because those projections had limited coverage. The integrated axis-level interpretation is summarized in **Table 1**, with the full evidence and claim checklist and cohort-level evidence summary provided in **Tables S12 and S13**.

**Table 1.**
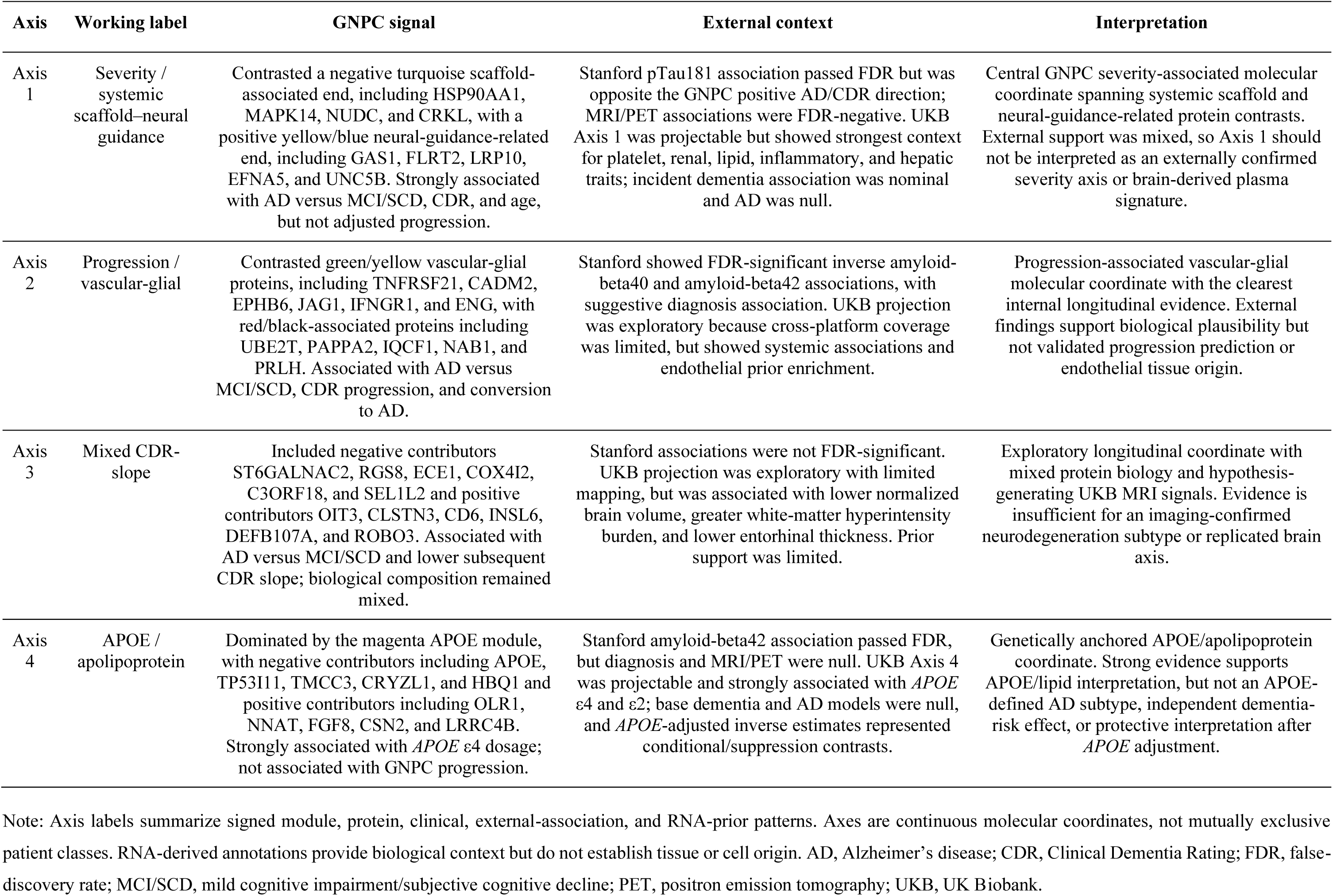
Integrated interpretation of continuous GNPC plasma proteomic axes.

## Discussion

In this multi-cohort plasma proteomic analysis, a brain-region- and cell-system-inspired hypothesis of Alzheimer’s disease (AD) subtyping was not supported as a set of discrete plasma proteomic classes. GNPC contained reproducible partitions, but the most stable solutions were weakly separated and added little clinical or external biomarker information after accounting for continuous module-derived axes. In contrast, four continuous axes captured most module-space variation and connected participant-level proteomic structure to diagnosis, CDR, progression, *APOE*, protein-level biology, and external molecular context. The main contribution of this study is therefore not a new AD subtype classifier, but a framework for representing plasma proteomic heterogeneity as continuous molecular coordinates.

This distinction is important for molecular phenotyping in AD. Prior transcriptomic and CSF proteomic studies have shown that AD is biologically heterogeneous and have motivated efforts to define molecular subtypes [6,7]. However, clustering-based categories can also arise from continuous biological gradients. In our data, repeated subsampling recovered a similar k=2 boundary, but participants remained densely distributed around that boundary and within-cluster heterogeneity was substantial. The k=2 solution was therefore stable without being clearly discrete. Because cluster assignment added negligible information beyond the four axes, treating those groups as subtypes would have imposed a categorical threshold without demonstrating a corresponding clinical or biomarker advantage. Stability alone is not sufficient for a subtype claim; separation, interpretability, incremental information, and coherent external support are also required [8,9].

The positive result of the study is the continuous axis framework. Continuous molecular coordinates preserve ordering, retain bidirectional protein contributions, and allow each participant to occupy several biological dimensions simultaneously. This is better aligned with the structure observed in GNPC than mutually exclusive subtype labels. Axis 1 captured a severity/systemic scaffold–neural guidance contrast and was strongly associated with AD versus MCI/SCD and CDR in GNPC, but its Stanford pTau181 association was directionally discordant likely due to Stanford ADRC small number of AD cases currently with a proportion of early-onset AD cases, as opposed to late-onset AD in GNPC. Axis 2 showed the clearest internal progression signal and carried vascular-glial annotation, but its Olink analogue was incompletely covered. Axis 3 had mixed biological composition despite clinical and exploratory MRI associations. Axis 4 was anchored to *APOE* and apolipoprotein biology, but its incident-disease interpretation changed after conditioning on *APOE* genotype. These features argue for graded, overlapping molecular states rather than discrete biological classes.

The external analyses defined the boundary of interpretation. Stanford ADRC provided clinically characterized fluid and imaging data. Four fluid biomarker associations passed FDR correction, yet the strongest Axis 1 association was opposite to the GNPC diagnosis and CDR direction, and while concordant MRI, amyloid PET, and tau PET associations did not survive modality-level correction. These findings do not negate the GNPC axes, but they prevent a simple severity-validation narrative. They also show the value of freezing axis direction before external testing: once direction is fixed, discordant biomarker evidence remains visible rather than being hidden by relabeling clusters or reversing post hoc interpretations.

UK Biobank provided a different form of evidence. Because the UKB analyses used Olink rather than SomaScan proteomics and were performed in a population-based cohort, they were designed as cross-platform contextualization rather than direct replication. The strongest UKB signals were systemic: platelet and blood-count traits, kidney function, lipids, inflammation, and liver measures. These associations were larger and more consistent than primary-axis cognition, MRI, or incident disease associations. This pattern is biologically informative because vascular, inflammatory, metabolic, hematologic, and organ-system biology may shape AD vulnerability, progression, or resilience. At the same time, the same systemic associations may also reflect aging, comorbidity, or general physiology rather than AD-specific mechanisms [10–13].

The UKB incident disease analyses were not sufficient to establish population risk markers. Axis 1 showed only a nominal association with all-cause dementia and no association with incident AD. Axis 4 was null in base dementia and AD models, and *APOE*-adjusted estimates changed direction. Because *APOE* is a dominant component and correlate of Axis 4, *APOE* genotype adjustment changes the estimand rather than merely removing confounding. These conditional results are useful precisely because they constrain interpretation: the projected axes should not be presented as dementia-risk predictors, and the study did not evaluate discrimination, calibration, decision utility, or clinically meaningful cut points.

Regional and cellular priors should also be interpreted cautiously. The study was motivated by regional and cellular vulnerability in AD, and RNA-derived priors helped annotate protein axes [3–5,14–17,22]. However, plasma proteins may reflect secretion, leakage, clearance, binding, peripheral expression, assay recognition, and systemic physiology. The presence of a brain-region or cell-type RNA enrichment among genes encoding measured proteins does not guarantee that the circulating protein abundance originated from that region or cell type. In the present study, neural annotations occurred alongside platelet, immune, endothelial, metabolic, and broadly expressed programs. The priors therefore support biological context, not claims of brain origin, tissue origin, or region-specific plasma subtypes.

Several limitations should be considered. GNPC axis derivation and internal clinical association were performed within the same broad discovery resource. Stanford ADRC used a previous Somalogic version and biomarker and imaging analyses were smaller than GNPC, particularly for tau PET, and Axis 1 showed unresolved directional discordance. SomaScan and Olink differ in target coverage, affinity reagents, epitope recognition, dynamic range, and analyte behavior; gene-level mapping preserves nomenclature but does not guarantee assay equivalence. Axis 2 and Axis 3 had limited Olink coverage and should be considered exploratory in UKB [25,26]. Finally, no axis has been prospectively calibrated for clinical use.

These limitations point to the next stage of validation. Independent longitudinal AD and MCI cohorts will be needed to determine whether the axes are reproducible, directionally stable, and clinically informative. The most informative designs would combine harmonized plasma proteomics with established AD biomarkers, including phosphorylated tau, neurodegeneration markers, amyloid and tau imaging, and structural MRI [23,24]. Same-platform replication, or prospectively defined cross-platform mapping, will be particularly important. Future studies should test whether continuous molecular coordinates improve prognosis, progression modeling, trial stratification, or treatment-response prediction beyond established demographic, genetic, clinical, and biomarker measures. Until such evidence is available, the axes should be viewed as research representations rather than clinical scores.

In summary, plasma proteomics did not identify robust discrete AD/MCI subtypes in this brain-region-and cell-system-inspired analysis. Instead, the data supported continuous molecular axes that captured neural, vascular-glial, APOE/apolipoprotein, and systemic components of plasma proteomic heterogeneity. Stanford ADRC cohort provided partial concordant external biomarker evidence, while UK Biobank contextualized the axes primarily through systemic physiology rather than brain specificity or dementia risk. These findings argue that plasma proteomic heterogeneity across the AD continuum is better represented by graded molecular coordinates than by hard subtype labels.

## Data Availability

GNPC data were accessed and analyzed through the Alzheimer's Disease Data Initiative AD Workbench subject to applicable data-use agreements. Stanford ADRC data are available through the Stanford ADRC under applicable institutional approvals and data-use restrictions. UK Biobank data are available to approved researchers through UK Biobank; analyses in this study were conducted under UK Biobank Application Number 45420. Derived summary results, supplementary tables, and figure-level outputs are provided with the manuscript and supplementary materials where permitted by the relevant data-use agreements.

https://addi-portal.alzheimersdata.org/

https://med.stanford.edu/adrc/researcher-resources.html

https://ams.ukbiobank.ac.uk/ams/

## Acknowledgments

We thank the participants, families, investigators, staff, and contributing cohorts of the Global Neurodegeneration Proteomics Consortium (GNPC) Stanford ADRC, and UK Biobank participants; whose biospecimens, clinical data, and proteomic data made this work possible. The GNPC harmonized dataset was accessed and analyzed through the Alzheimer’s Disease Data Initiative AD Workbench. Data discovery and/or analysis services contributing to this work were provided in-kind by the AD Data Initiative. We also acknowledge Gates Ventures and Johnson & Johnson for supporting the majority of GNPC biosample proteomic analyses, as described in the GNPC resource publication. UK Biobank data were analyzed on the Research Analysis Platform provided by DNANexus, and under Application Number 45420.

## Consent Statement

Each contributing cohort obtained approval from its local Institutional Review Board or Ethics Committee, and all participants gave written informed consent. Human studies were conducted in accordance with the Declaration of Helsinki. The current study protocol was granted an exemption by the Stanford University institutional review board because the analyses were carried out on deidentified data; therefore, additional informed consent was not required.

