## Supplementary_Figures for "Plasma proteomics reveals continuous molecular heterogeneity rather than discrete subtypes in Alzheimer’s disease"

This file contains supplementary figure captions and embedded supplementary figures. Supplementary table data are provided as a separate Excel workbook with one sheet per table.

### Supplementary Figures

**Figure S1. Expanded clustering diagnostics for GNPC discrete subtype tests.** Panels summarize selected clustering solutions, showing that reproducibility did not imply strong geometric separation or clinically useful discrete classes.

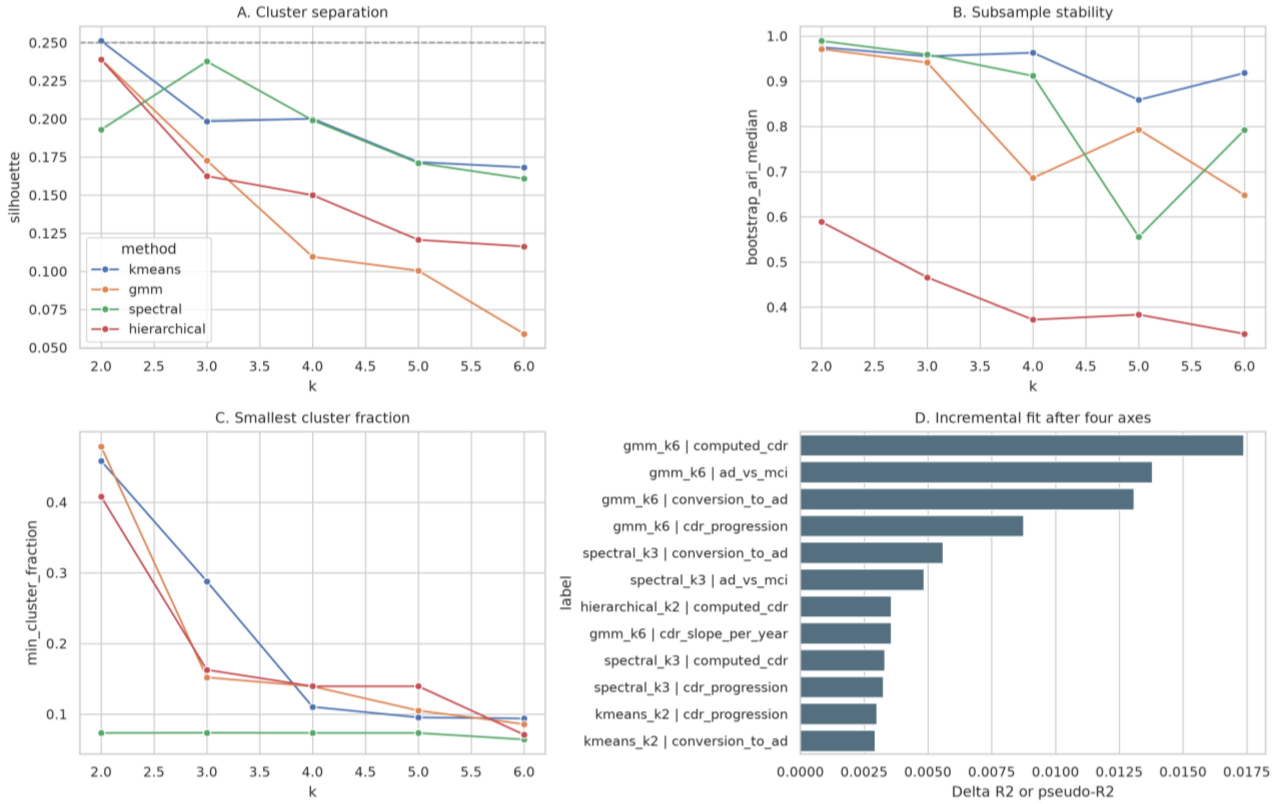

**Figure S2. Expanded protein and module heatmaps for the continuous GNPC axes**, emphasizing bidirectional protein contributions and module membership.

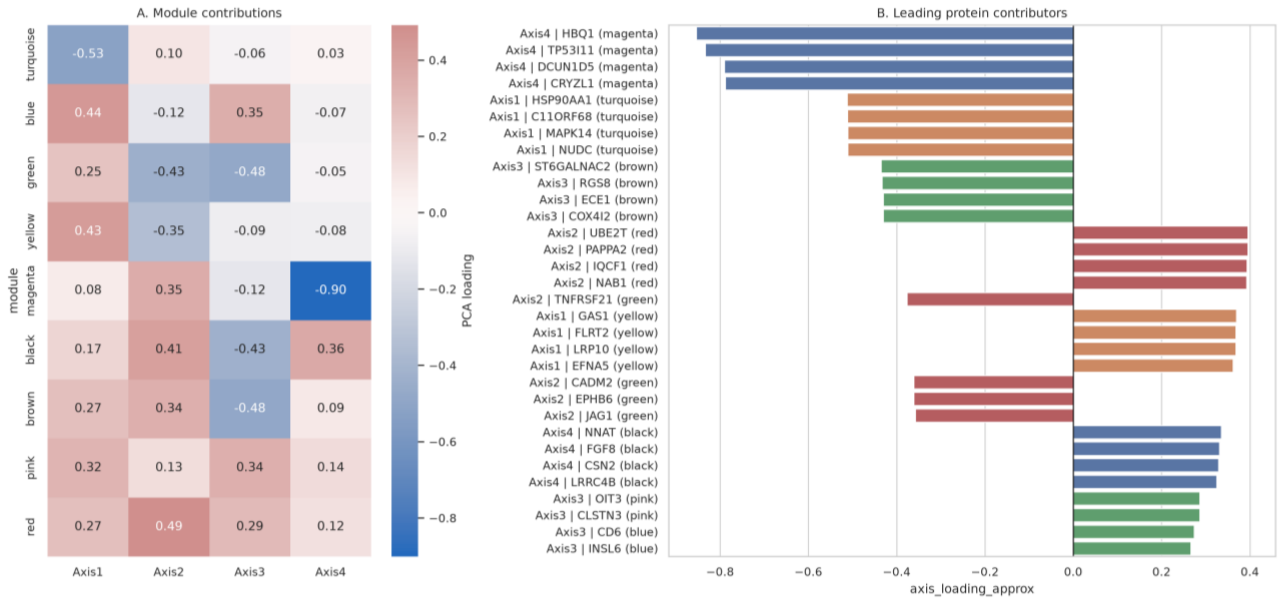

**Figure S3. Stanford external imaging and PET association panels. MRI thickness, amyloid PET, and tau PET.**

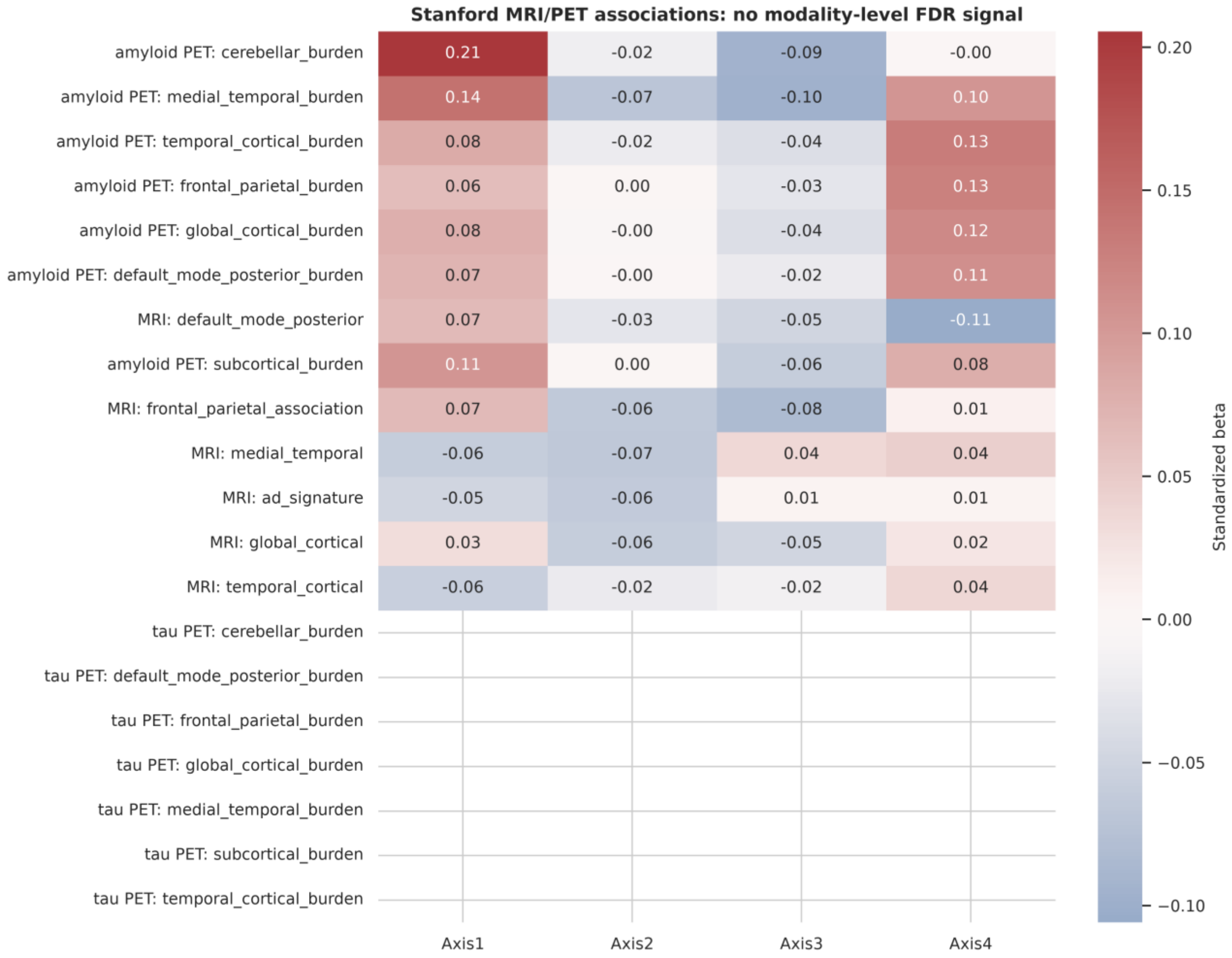

**Figure S4. Gene-level cross-platform projection coverage of GNPC axes in UK Biobank Olink.** Axis 1 and Axis 4 were primary projectable proxies; Axis 2 and Axis 3 had limited bidirectional coverage and are exploratory.

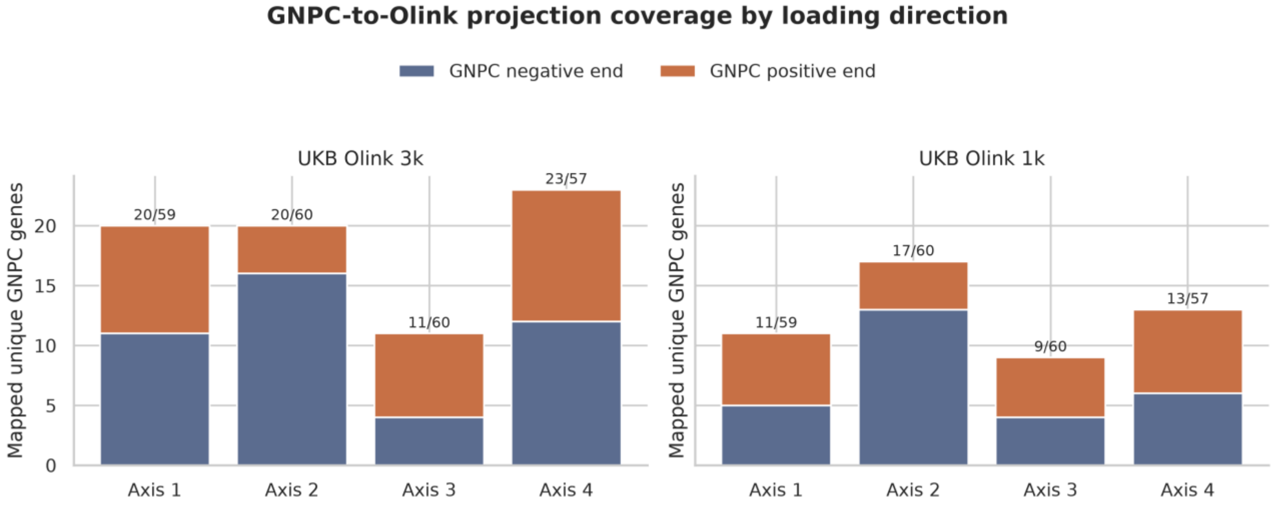


**Figure S5. Distributions of UK Biobank Olink proxy scores**, illustrating the continuous nature of projected axis scores rather than discrete patient classes.

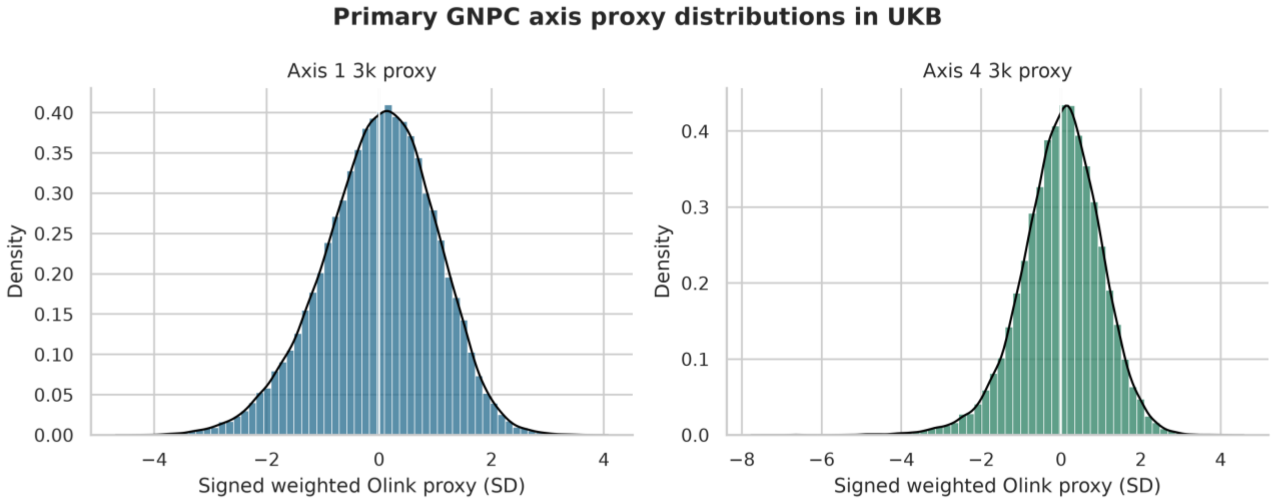

**Figure S6. UKB systemic trait heatmap.** UK Biobank intermediate phenotype associations with projected Olink axis scores. Strongest contextual signals involved systemic hematologic, renal, lipid, inflammatory, and hepatic traits.

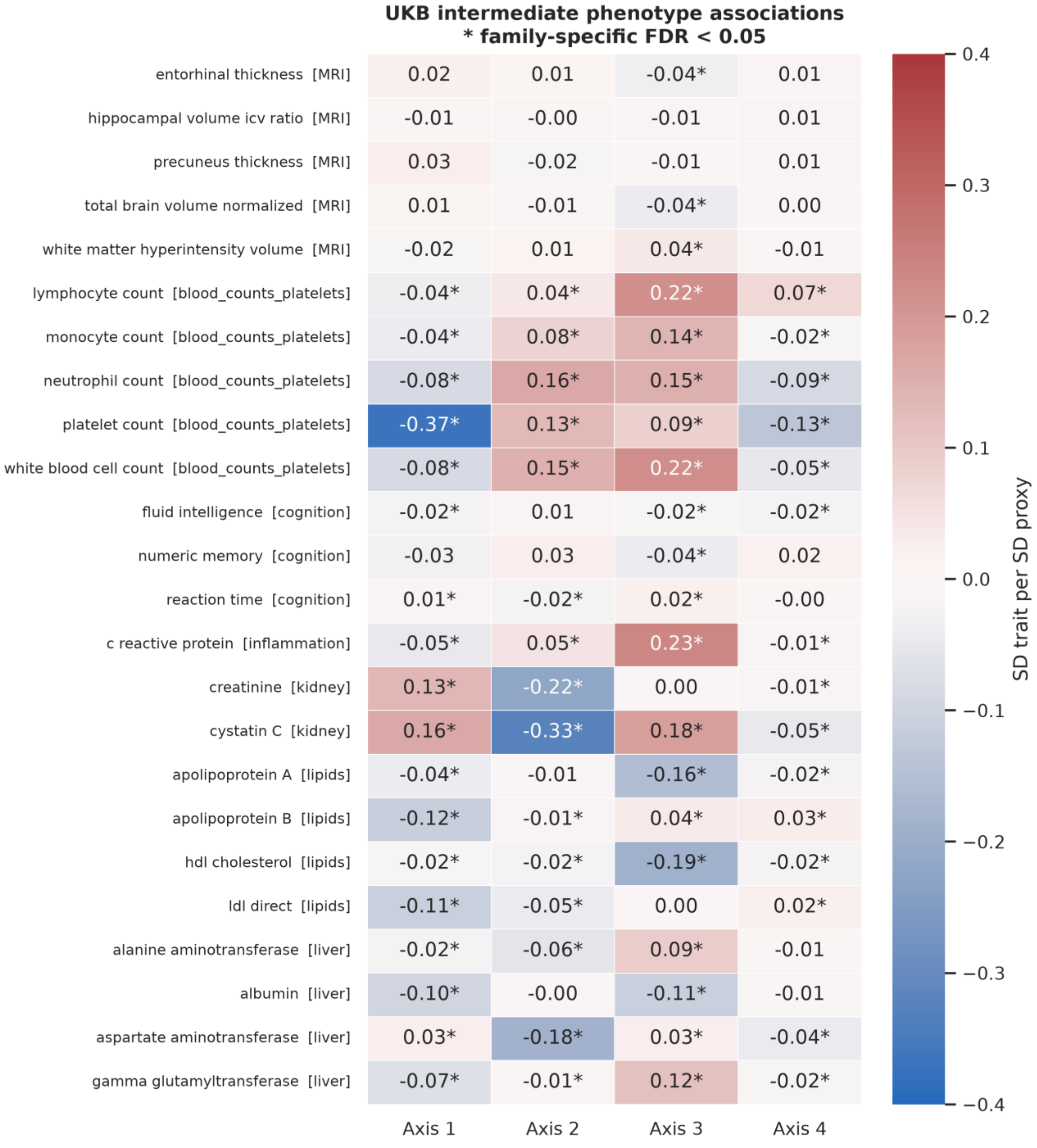

**Figure S7. UK Biobank axis cell/tissue prior enrichment.** Primary-axis set-level support was weak; Axis 2 endothelial and Axis 3 inhibitory-neuron findings are exploratory and do not establish tissue origin.

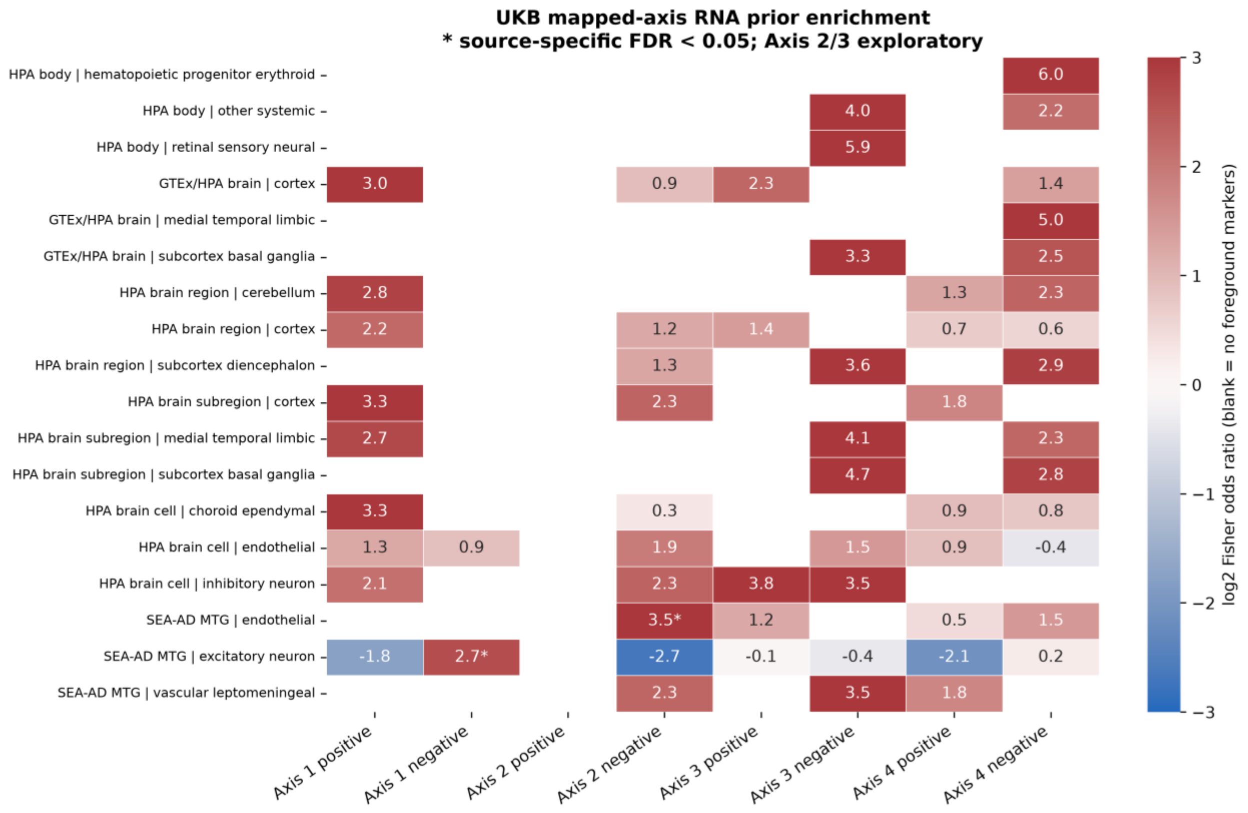

**Figure S8. Longitudinal stability of projected UKB Olink scores in the longitudinal samples with the Olink1k panel.** Intraclass correlations were moderate and were used as sensitivity/context rather than independent validation.

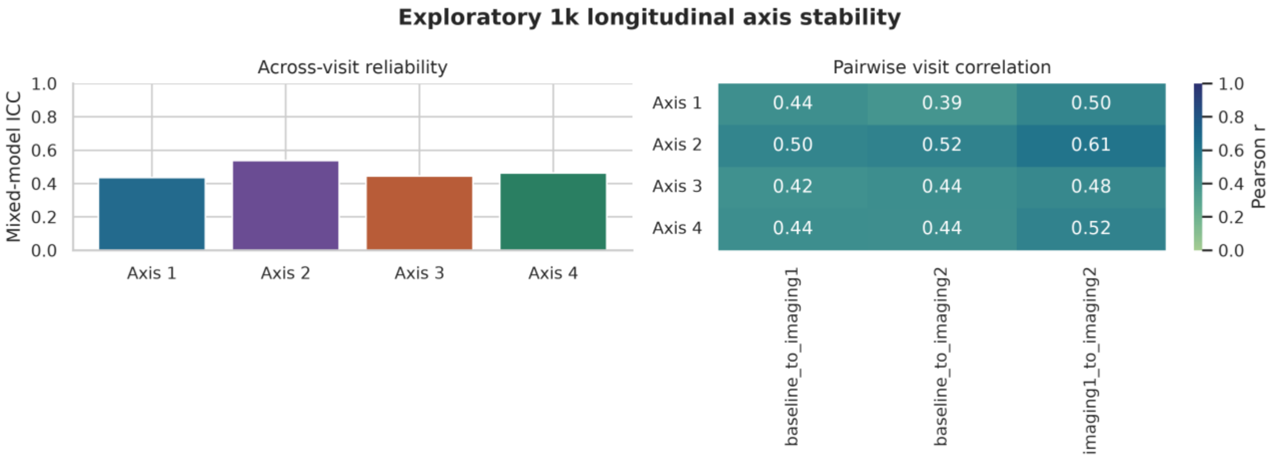
